# Lower Risk of Incident Cataracts and Diabetic Retinopathy amongst Individuals Treated with Sodium Glucose Cotransporter-2 Inhibitor Compared to Dipeptidyl Peptidase-4 Inhibitor in Type 2 Diabetes Mellitus

**DOI:** 10.1101/2024.03.25.24304828

**Authors:** Li Yen Goh, Oscar Hou In Chou, Sharen Lee, Teddy Tai Loy Lee, Jeremy Man To Hui, Hugo Pui Hok Him, Wing Tak Wong, Carlin Chang, Bernard Man Yung Cheung, Gary Tse, Jiandong Zhou

## Abstract

**Background/Aims:** Type 2 diabetes mellitus (T2DM) is an extremely prevalent disease with multisystem complications. We aim to compare the effects of two common glucose lowering medications; sodium glucose co-transporter 2 inhibitors (SGLT2I) and dipeptidyl peptidase-4 inhibitors (DPP4I), on the incidence of diabetic retinopathy and cataracts in T2DM patients in Hong Kong.

**Methods:** Retrospective population-based cohort study of T2DM patients treated with SGLT2I or DPP4I between 1^st^ January 2015 and 31^st^ December 2020. Propensity score matching (1:1 ratio) between SGLT2I and DPP4I users was performed on demographics, past co-morbidities, number of prior hospitalizations, duration from T2DM diagnosis to intial drug exposure, non-SGLT2I/DPP4I medications (including other anti-diabetes drugs), abbreviated modification of diet in renal disease, HbA1c, fasting glucose, and their time-weighted means. Sensitivity analysis using a one-year lag time and competing risk analyses using cause-specific and sub-distribution hazard models were conducted.

**Results:** This study cohort included 26 165 SGLT2I and 42 796 DPP4I users (total: N=68 961 patients; 56.43% males, median age: 62.0 years old (standard deviation (SD): 12.8)). Over a median follow-up of 5.56 years (IQR: 5.24-5.80) and after propensity score matching (SGLT2I: N=26 165; DPP4I: N=26 165), SGLT2I users had lower incidences of cataract (4.54% vs. 6.64%%, standardised mean difference [SMD]=0.09) and diabetic retinopathy (3.65 vs. 6.19, SMD=0.12) compared to DPP4I users. SGLT2I use was associated with lower risks of new onset cataract (HR: 0.67, 95% CI: [0.62– 0.72] P<0.0001) and diabetic retinopathy (hazard ratio [HR]: 0.57, 95% confidence interval [CI]: [0.53–0.62], P<0.0001). These associations remained significant on multivariable Cox regression ;cataract: HR: 0.69, 95% CI: 0.64–0.75 (P<0.0001); diabetic retinopathy: HR: 0.68, 95% CI: 0.63–0.75 (P<0.0001).

**Conclusions:** Amongst T2DM patients in Hong Kong, SGLT2I use was associated with lower risks of new onset cataract or diabetic retinopathy compared to DPP4I use.

**Synopsis/Precis:** Sodium glucose cotransporter-2 inhibitor (SGLT2I) use was associated with lower rates of new onset diabetic retinopathy and cataracts compared to dipeptidyl peptidase-4 inhibitor (DPP4I) use in patients with type 2 diabetes melllitus (T2DM) from Hong Kong.

**What is already known on this topic:** Various glucose lowering medications may have additional beneficial or aggravating properties for/against diabetic retinopathy and cataract formation in diabetic populations beyond their glucose lowering capabilities.

**What this study adds:** This study showed that SGLT2I use was associated with significantly lower rates of new onset cataracts and diabetic retinopathy when compared to DPP4I use in a T2DM population in Hong Kong. Additionally, to the best of our knowledge, this is the first population-based study on the effects of SGLT2I and DPP4I use on the development of cataracts in individuals with T2DM.

**How this study might affect research, practice or policy:** This study provides preliminary data for further evaluation of SGLT2I and DPP4I use in preventing the incidence and progression of cataracts and diabetic retinopathy in a T2DM individuals. This study may also aid clinicians in deciding between SGLT2 and DPP4I if microvascular retinal complications and cataracts are a concern in individual cases.

## Introduction

Type 2 diabetes mellitus (T2DM) is a widespread disease with increasing prevalence globally. It is predicted that by 2030, up to 439 million people worldwide will have T2DM (1). The potential multi-system morbidities and mortality associated with this disease are well documented. Cataracts and diabetic retinopathy are recognised complications, with sight threatening capabilities; both feature among the top five causes of global blindness (2). It is estimated that by 2030, 191 million individuals with T2DM will have diabetic retinopathy (2). There have been several historic landmark trials which have suggested that intensive blood glucose control (HbA1c <7%) achieved through medication is associated with lower risks of microvascular diabetic complications (3–6). Additionally, patients with T2DM are at higher risk of developing cataracts at an earlier age than the general population (7).

Metformin is the commonest first-line anti-diabetic agent, and if glycemic control is not optimally achieved, agents such as dipeptidyl-peptidase 4 inhibitors (DPP4I), pioglitazone and sulphonylureas or dual therapy with sodium-glucose transport protein 2 inhibitors (SGLT2I) can be prescribed. Additionally, the National Institute for Health and Care Excellence (NICE) guidance has recently advised that DPP4I or SGLT2I are among the recommended first-line treatments if metformin is contraindicated or not tolerated (8).

### Aims

There have been arguments that various glucose lowering agents may have additional protective or aggravating properties towards ocular microvascular complications and cataract formation beyond their glucose lowering capabilities (7, 9, 10). This study aims to compare the outcomes of incident diabetic retinopathy and cataract development between SGLT2I and DPP4I users in a T2DM population in Hong Kong.

### Objectives

**1.** To compare the outcomes of new-onset cataracts between SGLT2I and DPP4I users in a T2DM population in Hong Kong identified through the Clinical Data Analysis and Reporting System (CDARS), a city-wide database that centralises patient information and using the International Classification of Diseases, Ninth Revision, Clinical Modification (ICD-9) codes, National Centre of Health Statistics, Centers for Disease Control and Prevention (11)
**2.** To compare the outcomes of new-onset diabetic retinopathy between SGLT2I and DPP4I users in a T2DM population in Hong Kong identified through the CDARS, using ICD-9 codes, National Centre of Health Statistics, Centers for Disease Control and Prevention (11)

## Materials and Methods

### Study design and population

This study obtained ethics approval from The Joint Chinese University of Hong Kong–New Territories East Cluster Clinical Research Ethics Committee and The Institutional Review Board of the University of Hong Kong/Hospital Authority Hong Kong West Cluster. This was a retrospective, territory-wide cohort study of T2DM patients with SGLT2I or DPP4I use between approximately 1^st^ January 2015 and 31^st^ December 2020 in Hong Kong. The patients were identified via ICD-9 codes from the Clinical Data Analysis and Reporting System (CDARS), a city-wide database that centralises patient information from 43 public hospitals or their associated ambulatory or outpatient facilities managed by the Hong Kong Hospital Authority to establish comprehensive medical data, including clinical characteristics, laboratory results, drug treatments and disease diagnosis. CDARS has been previously utilised by our team and other teams in Hong Kong (12, 13). ICD-9 codes definitions were obtained from the Centers for Disease Prevention and Control website (11).

Patients with SGLT2I or DPP4I use during the study period were identified (76 147). Excluded patients: without complete demographics (N=17), under 18 years old (N=108), with prior diagnoses of cataracts, glaucoma, congenital anomalies/dystrophies, rubeosis iridis, ocuar trauma/inflammation, diabetic eyes disease, retinal vascular occlusion, age related macular degeneration, ocular malignancy, other proliferative retinopathy or severe eye events (N = 4693), with prior use of setroids in the eye (N = 1 302) and those who had prebious procedure in the eyes for diabetic retinopathy diabetic macular oedema, anti-vascular endothelial growth factor injections (N = 1 066). Individuals who took medications which could cause retinopathy; anti-malarials/anti-rheumatics including, quinine, hydroxychloroquine, mefloquine; anti-psychotics including phenothiazines (fluphenazine, promazine, thirodazine, mesoridazine), prochlorperazine, chlorpromazine, perphenazine; non-steroidal anti-inflammatory drugs, including indomethacin, antibiotics including ethambutol; heavy metal antagonists, including desferrioxamine and anti-estrogen including tamoxifen were also excluded (.

Clinical and biochemical data were extracted for the present study. Patient demographics include sex and age at commencement of initial SGLT2I/DPP4I use. Outcomes of new-onset cataracts and diabetic retinopathy were extracted if the condition was documented in either one or both eyes, based on ICD-9 coding. Prior co-morbidities were also extracted, including diabetes with chronic complications, diabetes without chronic complications, gout, heart failure, hyperlipidaemia, hypertension, hypoglycemia and hyperglycaemia (using hospitalisations as a proxy measure), ischemic heart disease, liver diseases, acute myocardial infarction, peripheral vascular disease, renal diseases, stroke/transient ischemic attack, VT/VF/SCD (VT: ventricular tachycardia, VF: ventricular fibrillation, SCD: sudden cardiac death), anaemia, high Body Mass Index (BMI), and cancer, based on ICD-9 codes. Charlson’s standard comorbidity index was also calculated. Mortality was recorded using the International Classification of Diseases Tenth Edition (ICD-10) coding (14).

Medication histories were also extracted, including the use of metformin, sulphonylurea, insulin, acarbose, thiazolidinedione, glucagon-like peptide-1 receptor agonists, statins and fibrates. Baseline laboratory data, including complete blood count, biochemical tests, glycemic and lipid profiles were extracted. In addition, the abbreviated modification of diet in renal disease (AMDRD) equation was calculated to assess the renal function of the included patients (15).

### Outcomes and statistical analysis

The study outcomes were new onset cataract and diabetic retinopathy. Mortality data were obtained from the Hong Kong Death Registry, a population-based official government registry with the registered death records of all Hong Kong citizens linked to CDARS. The follow-up period was 31^st^ December 2020 or until death, whichever was earlier.

Descriptive statistics were used to summarise the baseline, clinical and biochemical characteristics of patients with SGLT2I and DPP4I treatments. For baseline clinical characteristics, continuous variables were presented as median (95% confidence interval [CI]/ interquartile range [IQR]) or mean (standard deviation [SD]) and categorical variables were presented as total number (percentage). Continuous variables were compared using the two-tailed Mann-Whitney U test, whilst the two-tailed Chi-square test with Yates’ correction was used to test 2 × 2 contingency data.

Propensity score matching with 1:1 ratio between SGLT2I and DPP4I users based on demographics, prior co-morbidities and hospitalisations, AMDRD, HBA1c, fasting glucose, use of different medication classes (including other anti-diabetic drugs) and their time-weighted means, were performed using the nearest neighbor search strategy with the caliper as 0.1. Univariable and multivariable Cox regression models were used to identify significant risk predictors for the study outcomes. Competing risk analysis models (cause-specific and sub-distribution) were considered with mortality outcomes. A standardised mean difference (SMD) of less than 0.2 between the treatment groups post-weighting was considered an adequate balance. The hazard ratio (HR), 95% CI and P-value were reported. Statistical significance is defined as P-value < 0.05. Statistical analyses were performed with RStudio software (Version: 1.1.456) and Python (Version: 3.6). Stata software (Version 13.0) was used for propensity score matching.

## Results

### Basic characteristics of the study cohort

This study cohort included 68 961 patients (56.43% males, median age: 62.0 years old (standard deviation (SD) 12.8) with 26 165 SGLT2I users and 42 796 DPP4I users (**Figure 1**). The baseline and clinical characteristics of SGLT2I and DPP4I users before and after 1:1 propensity score matching are shown in **Table 1**. After a median follow-up duration of 5.56 years (IQR: 5.24 - 5.8), in the propensity score-matched cohort (SGLT2I: N=26 165; DPP4I: N=26 165), 2 928 patients (5.59%) developed new onset cataract and 2 577 (4.92%) developed diabetic retinopathy (**Table 2**). SGLT2I users had lower incidences of new onset cataract, 1 190 (4.54%) vs. 1 738 (6.64%), standardized mean difference [SMD]=0.09 and new onset diabetic retinopathy 956 (3.65%) vs. 1 621 (6.19%), SMD=0.12),compared to DPP4I users (**Table 2**). The time to new onset cataracts was similar for both groups (**Table 2**), the mean time in years for SGLT2I was 5.4 years (SD: 0.8) and 5.2 years (SD: 1.1) with a SMD of 0.21. The time to new onset diabetic diabetic retinopathy was also similar for both groups (**Table 2**), the mean time in years for SGLT2I was 5.4 years (SD: 0.8) and 5.2 years (SD: 1.1) with a SMD of 0.23.

**Figure 1.**
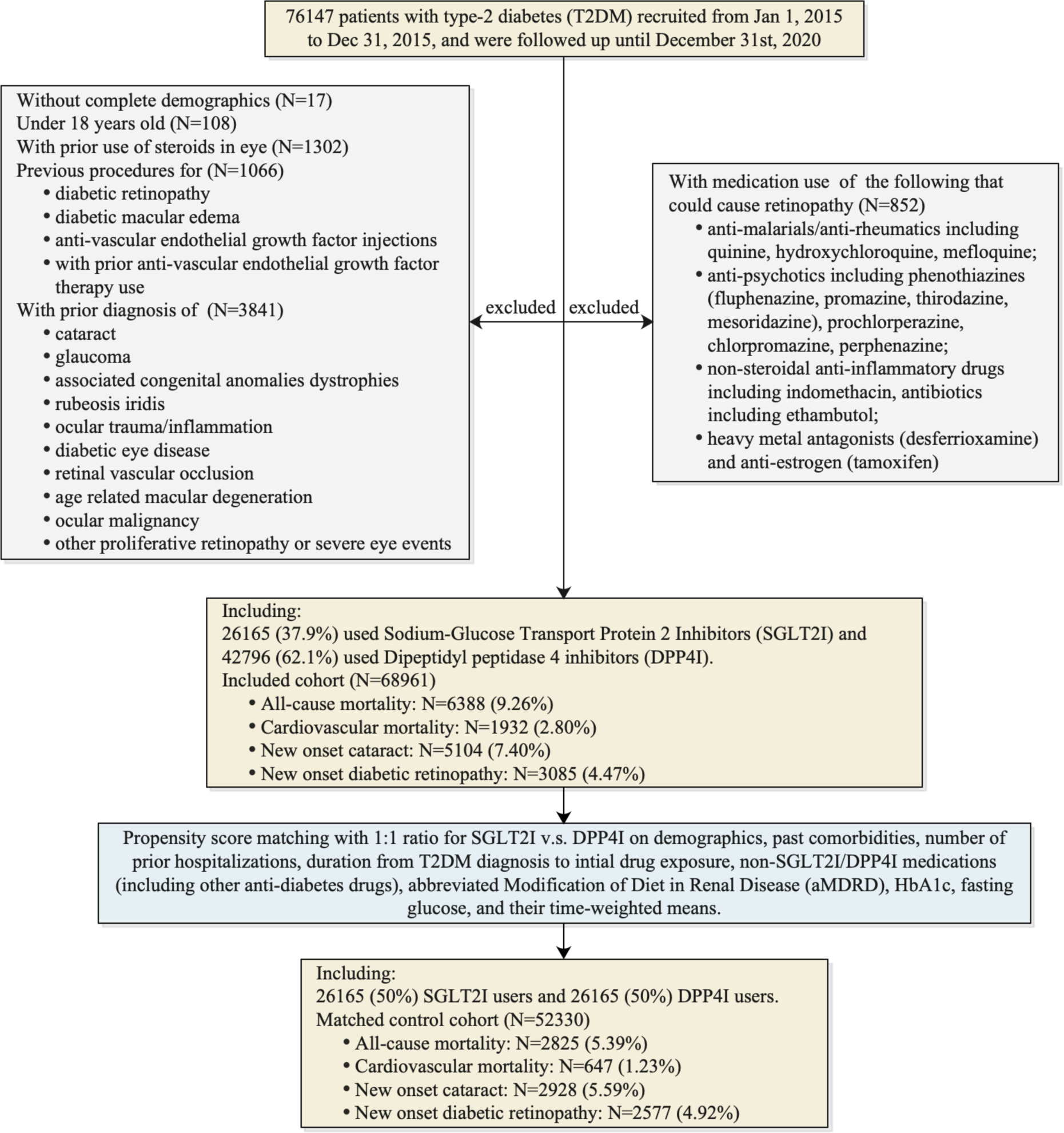
Flowchart of data processing. SGLT2I: sodium glucose cotransporter-2 inhibitor; DPP4I: dipeptidyl peptidase-4 inhibitor.

**Table 1.**
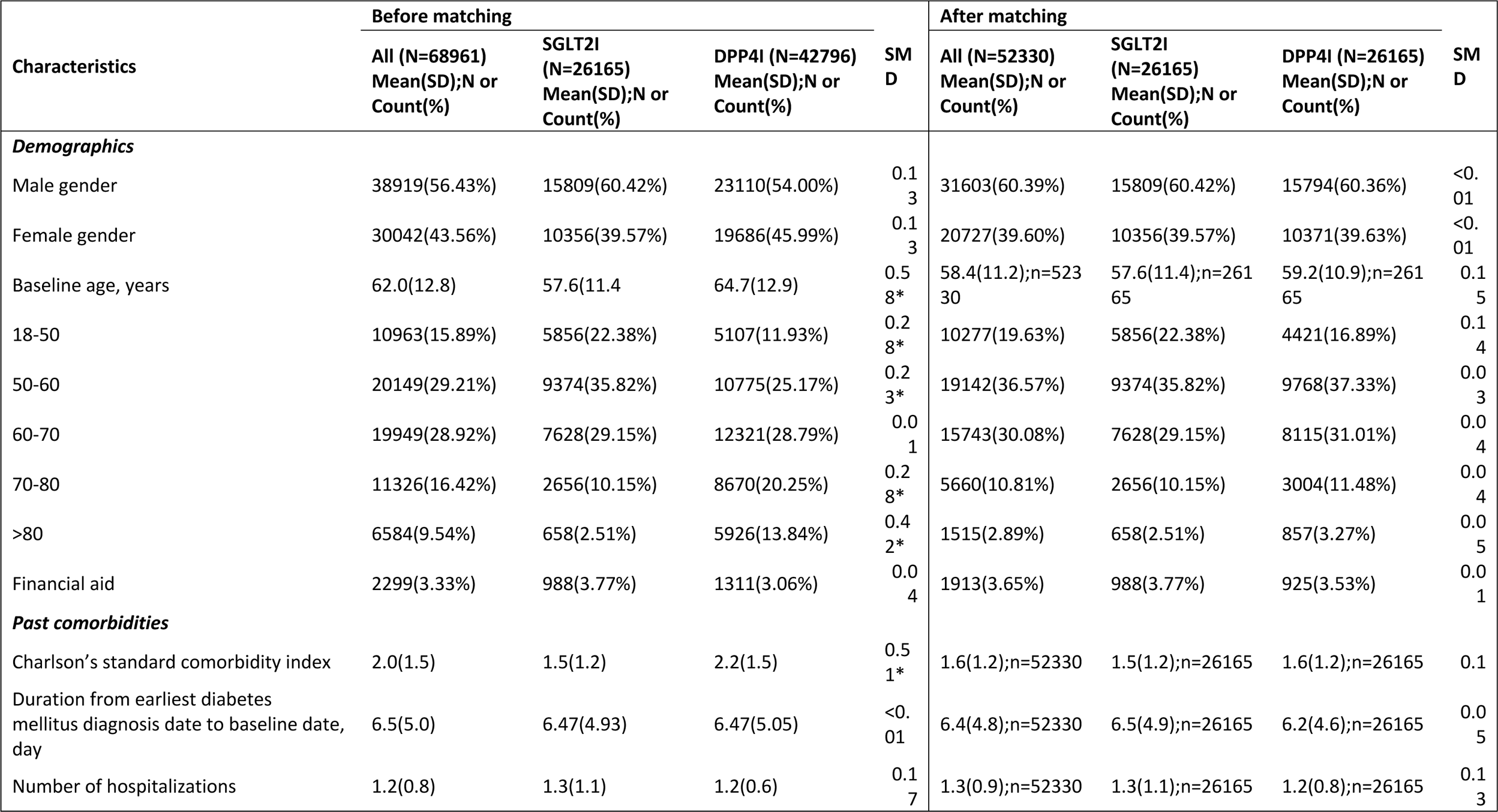
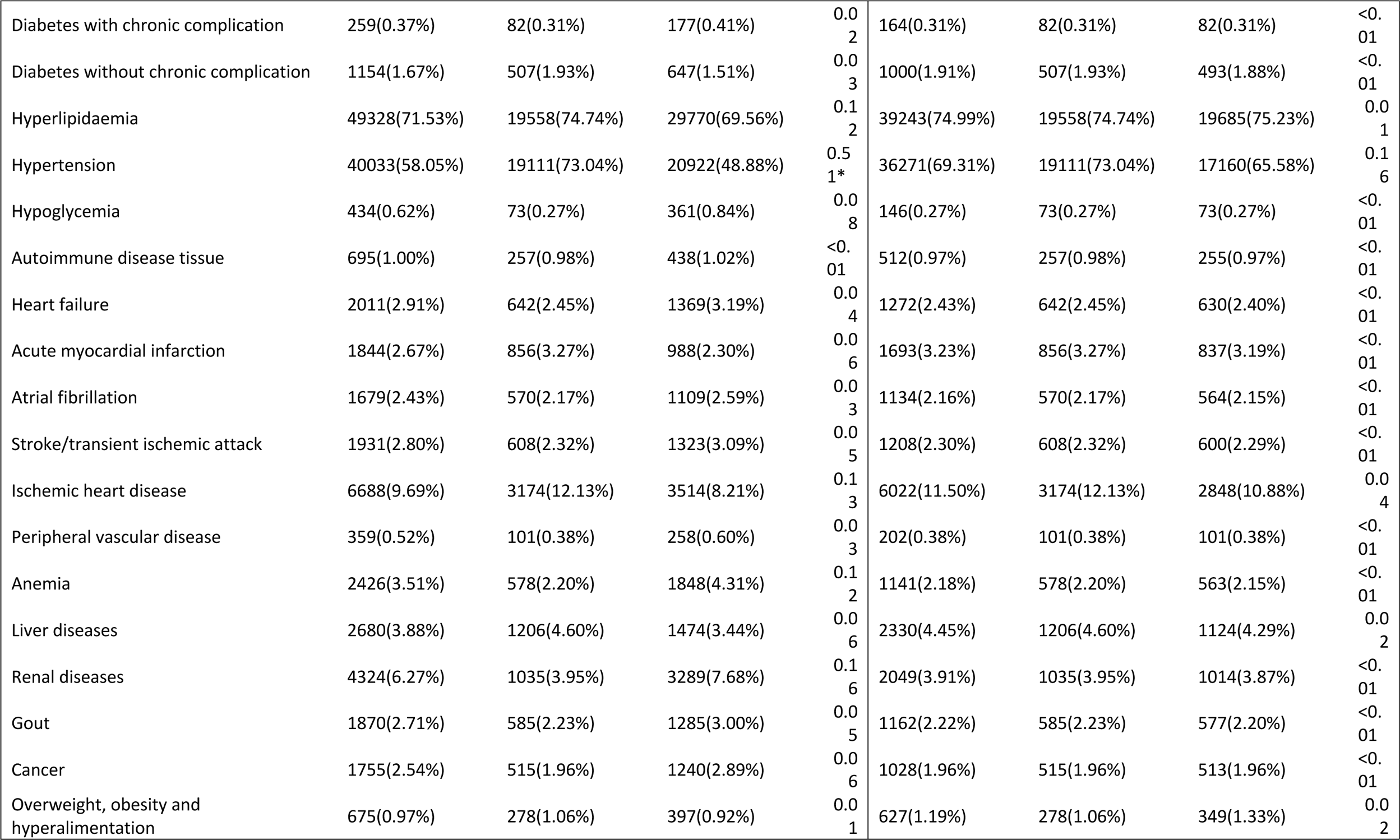
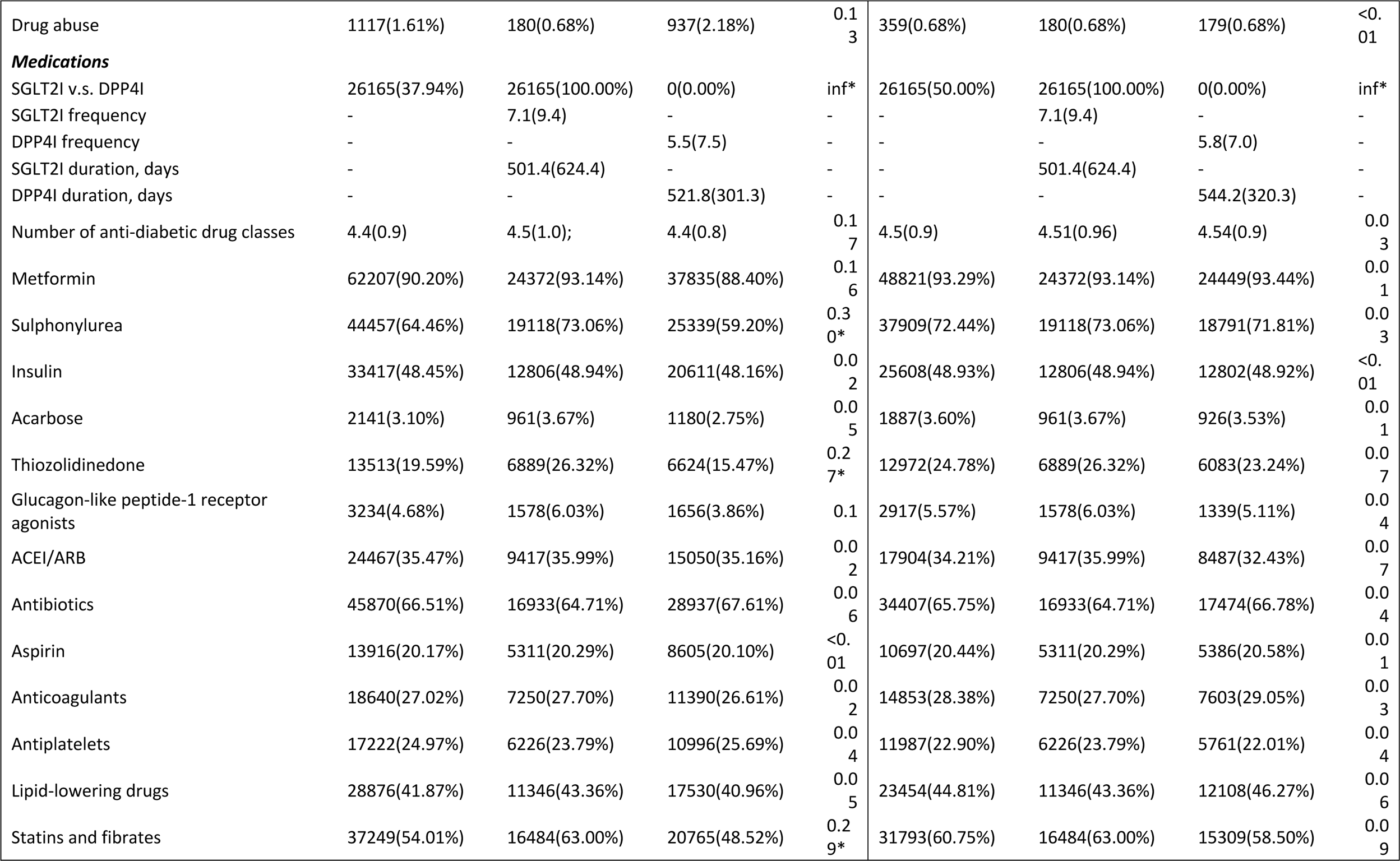
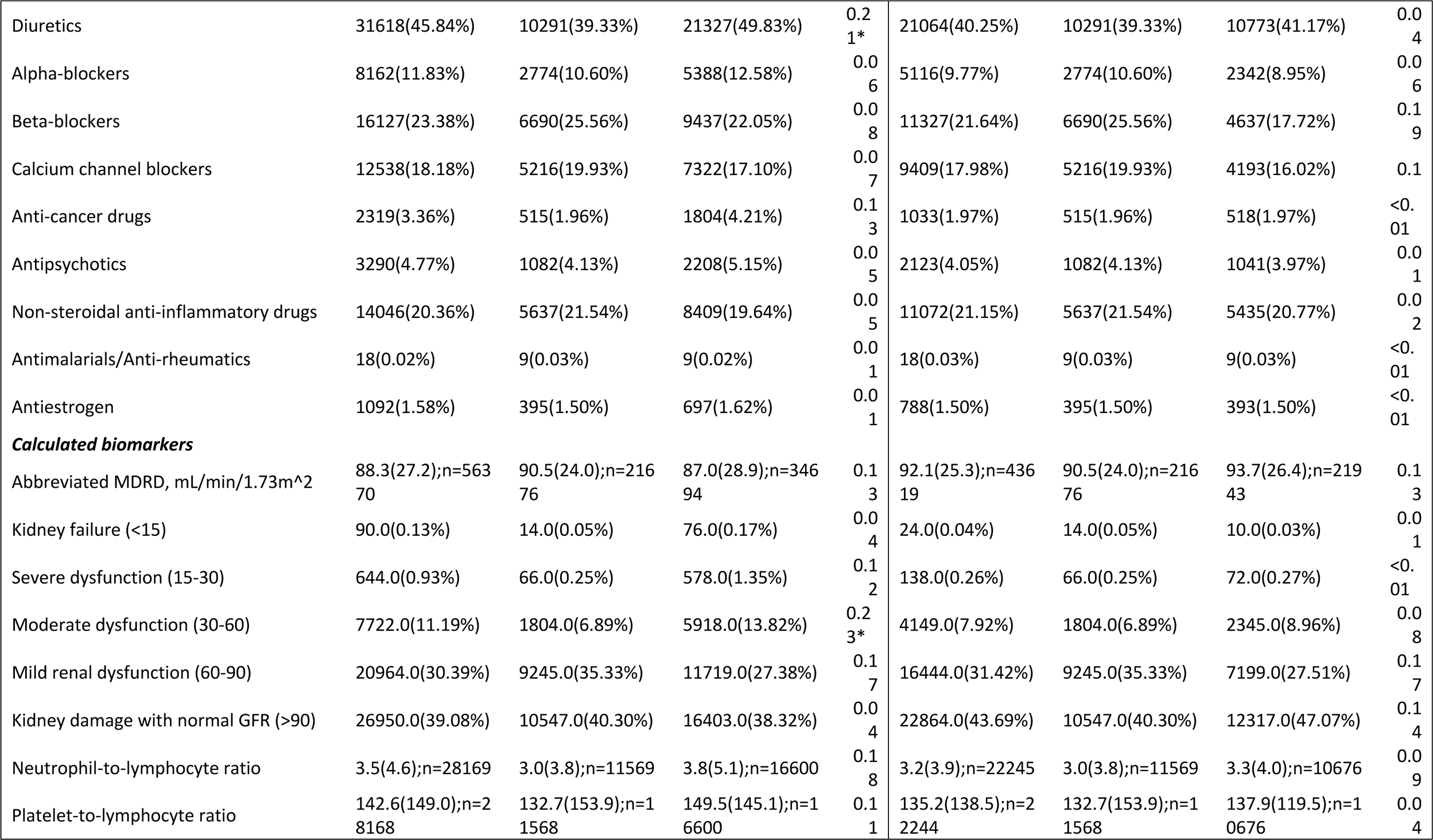
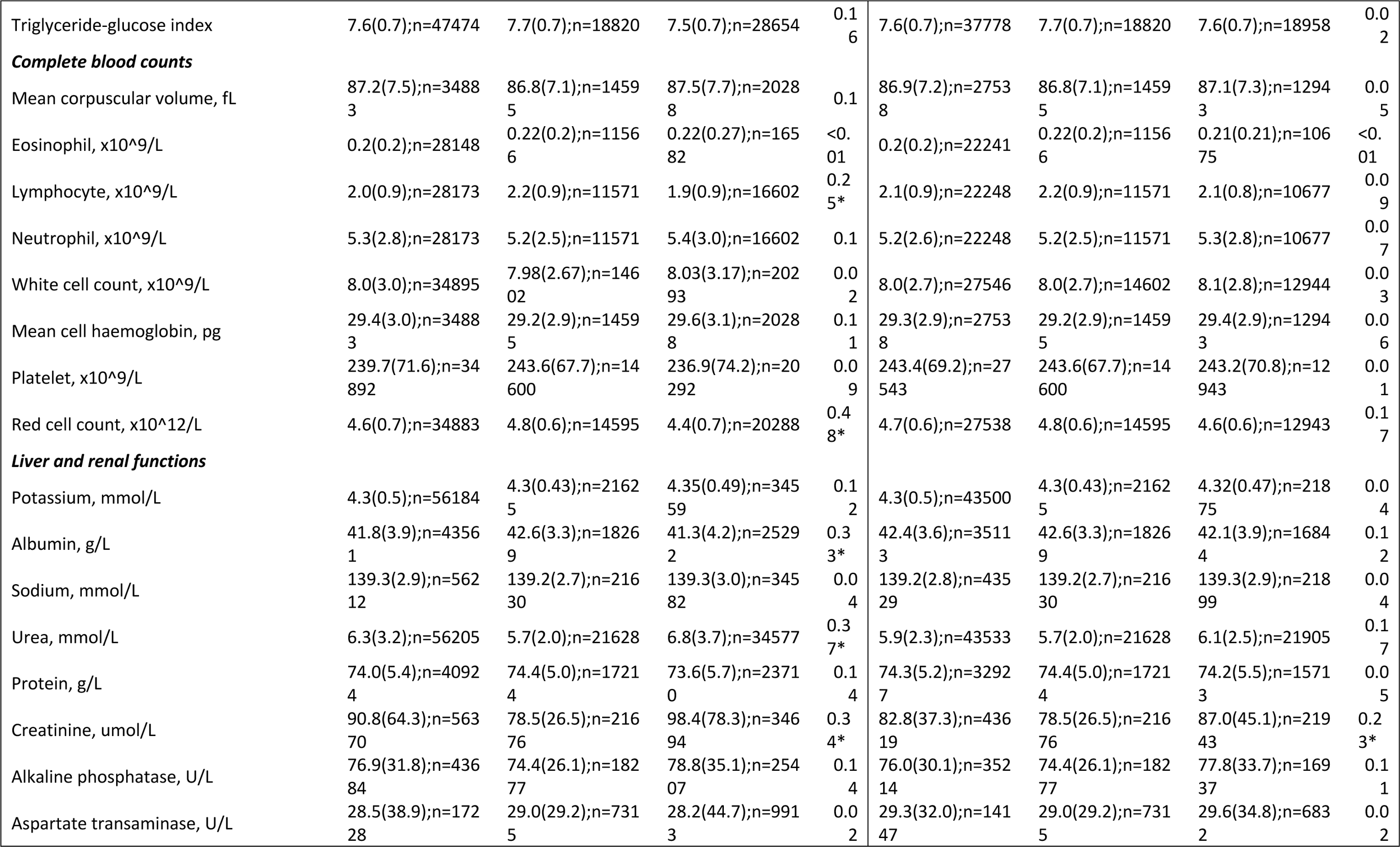
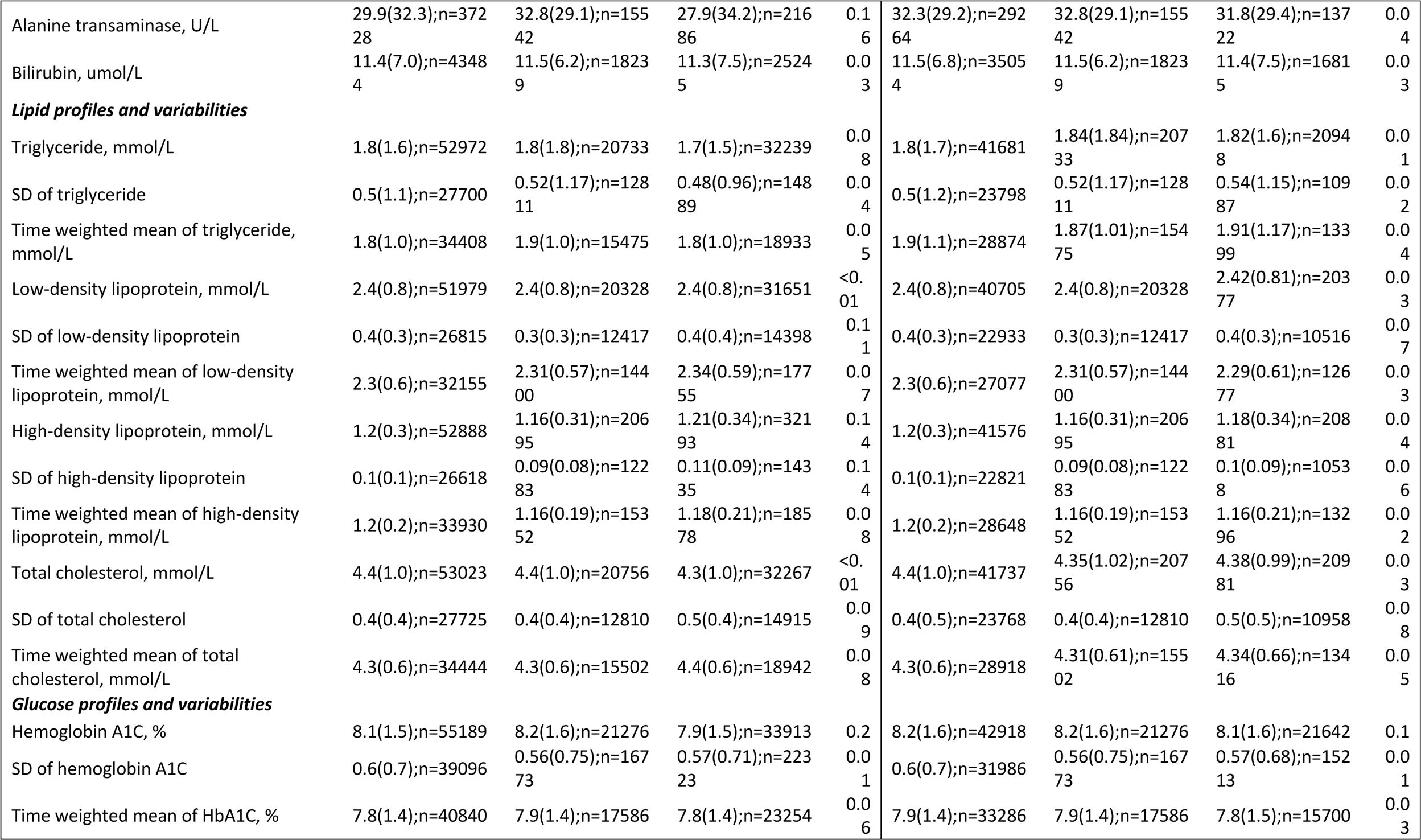
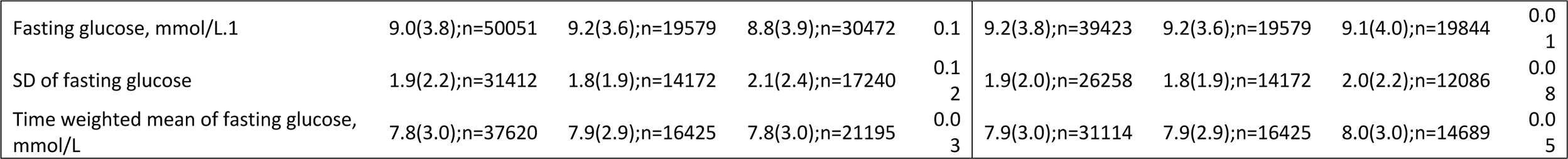
Baseline and clinical characteristics of patients with SGLT2I v.s. DPP4I use before and after propensity score matching (1:1). * for SMD≥0.1; SGLT2I: sodium glucose cotransporter-2 inhibitor; DPP4I: dipeptidyl peptidase-4 inhibitor; SD: standard deviation; CV: Coefficient of variation; MDRD: modification of diet in renal disease; ACEI: angiotensin-converting enzyme inhibitors; ARB: angiotensin II receptor blockers.

**Table 2.** Outcome characteristics in patients by SGLT2I v.s. DPP4I use before and after propensity score matching (1:1). * for SMD≥0.1; SGLT2I: sodium glucose cotransporter-2 inhibitor; DPP4I: dipeptidyl peptidase-4 inhibitor.

| Characteristics | Before matching |  |  |  | After matching |  |  |  |
| --- | --- | --- | --- | --- | --- | --- | --- | --- |
|  | All (N=68961)<br>Mean(SD); N or<br>Count(%) | SGLT2I<br>(N=26165)<br>Mean(SD);N or<br>Count(%) | DPP4I (N=42796)<br>Mean(SD); N or Count(%) | SMD | All (N=52330)<br>Mean(SD);N or<br>Count(%) | SGLT2I<br>(N=26165)<br>Mean(SD); N or<br>Count(%) | DPP4I (N=26165)<br>Mean(SD); N or Count(%) | SMD |
| All-cause mortality | 6388(9.26%) | 727(2.77%) | 5661(13.22%) | 0.39* | 2825(5.39%) | 727(2.77%) | 2098(8.01%) | 0.23* |
| Time to mortality, years | 5.4(0.7) | 5.5(0.4) | 5.3(0.9) | 0.38* | 5.5(0.6) | 5.5(0.4) | 5.4(0.7) | 0.26* |
| Cardiovascular mortality | 1932(2.80%) | 148(0.56%) | 1784(4.16%) | 0.24* | 647(1.23%) | 148(0.56%) | 499(1.90%) | 0.12 |
| Time to CAD mortality, years | 5.4(0.7) | 5.5(0.4) | 5.3(0.9) | 0.38* | 5.5(0.6) | 5.5(0.4) | 5.4(0.7) | 0.26* |
| New onset cataract | 5104(7.40%) | 1190(4.54%) | 3914(9.14%) | 0.18 | 2928 (5.59%) | 1190 (4.54%) | 1738 (6.64%) | 0.09 |
| Time to cataract, year | 5.1(1.2) | 5.4(0.8) | 5.0(1.3) | 0.36* | 5.3(1.0) | 5.4(0.8) | 5.2(1.1) | 0.21* |
| New onset diabetic retinopathy | 3085(4.47%) | 956(3.65%) | 2129(4.97%) | 0.07 | 2577(4.92%) | 956(3.65%) | 1621(6.19%) | 0.12 |
| Time to Diabetic retinopathy, year | 5.2(1.0) | 5.4(0.8) | 5.1(1.1) | 0.3* | 5.3(1.0) | 5.4(0.8) | 5.2(1.1) | 0.23* |

### Univariable, multivariable Cox regression models

The detailed results of univariable Cox regression in the unmatched and matched cohorts are shown in **Table 3** and 4. In the matched cohort, compared to DPP4I users, SGLT2I users showed lower risks of new onset cataract (HR: 0.67, 95% CI: [0.62–0.72] P<0.0001) and new onset diabetic retinopathy (HR: 0.57, 95% CI: [0.53–0.62], P<0.0001) (**Table 3**). After adjustments for significant demographics, co-morbidities as listed above, length of T2 diabetes mellitus, duration of usage of SGLT2I or DPP4I use, non-SGLT2I/DPP4I diabetic medications, laboratory markers (lipid and glucose markers) and including sensitivity analyses for chronic kidney disease stage 4 and 5, peritoneal dialysis and haemodialysis, as well as previous use of insulin and glucagon-like peptide-1 receptor agonists (GLP1-RA). SGLT2I users had a statistically significantly lower association with new onset cataract (HR: 0.76, 95% CI: [0.67–0.87 P<0.0001) and new onset diabetic retinopathy (HR:0.51, 95% CI: [0.42 – 0.61, P <0.0001) (**Table 4**). This startistically significantly lower association with new cataracts was present for common SGLT2I medications including canagliflozin and ertugliflozin, but not dapafliflozin and empagliflozin. This statistically significantly lower association with new onset diabetic retinopathy was present for the common SGLT2I medication dapagliflozin but not canagliflozin, ertugliflozin and empagliflozin (**Table 4**).

**Table 3.** Univariate Cox regression models in the match cohort. * for SMD≥0.1; HR: hazard ratio; CI: confidence interval; SGLT2I: sodium glucose cotransporter-2 inhibitor; DPP4I: dipeptidyl peptidase-4 inhibitor.

|  | New onset cataract<br>HR [95% CI];P value | New onset diabetic retinopathy<br>HR [95% CI];P value |
| --- | --- | --- |
| SGLT2I v.s. DPP4I | 0.67[0.62-0.72];<0.0001*** | 0.57[0.53-0.62];<0.0001*** |
| Dapagliflozin v.s. DPP4I | 0.77[0.71-0.84];<0.0001*** | 0.63[0.57-0.69];<0.0001*** |
| Empagliflozin v.s. DPP4I | 0.76[0.67-0.87];<0.0001*** | 0.73[0.64-0.84];<0.0001*** |
| Canagliflozin v.s. DPP4I | 0.68[0.60-0.78];<0.0001*** | 0.72[0.63-0.83];<0.0001*** |
| Ertugliflozin v.s. DPP4I | 0.68[0.56-0.82];<0.0001*** | 0.69[0.56-0.84];0.0002*** |

**Table 4.**
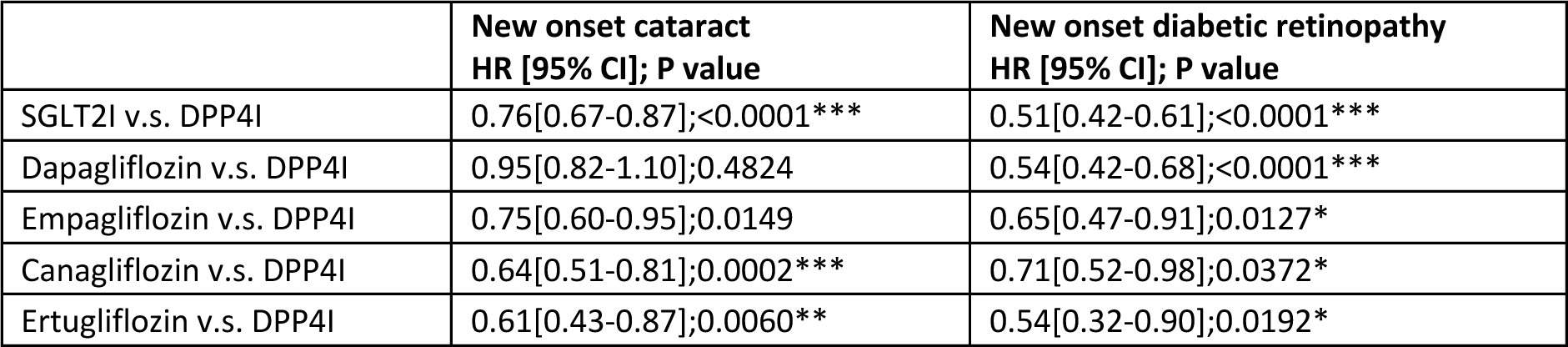
Cox univariate sensitivity analysis with exclusion of patient’s chronic kidney disease stage 4 or 5 (eGFR <30), peritoneal dialysis or haemodialysis and patients who used insulin and/or GLP-1 RA in the matched cohort. * for p≤ 0.05, ** for p ≤ 0.01, *** for p ≤ 0.001; HR: hazard ratio; CI: confidence interval; SGLT2I: sodium glucose cotransporter-2 inhibitor; DPP4I: dipeptidyl peptidase-4 inhibitor.

|  | New onset cataract<br>HR [95% CI]; P value | New onset diabetic retinopathy<br>HR [95% CI]; P value |
| --- | --- | --- |
| SGLT2I v.s. DPP4I | 0.76[0.67-0.87];<0.0001*** | 0.51[0.42-0.61];<0.0001*** |
| Dapagliflozin v.s. DPP4I | 0.95[0.82-1.10];0.4824 | 0.54[0.42-0.68];<0.0001*** |
| Empagliflozin v.s. DPP4I | 0.75[0.60-0.95];0.0149 | 0.65[0.47-0.91];0.0127* |
| Canagliflozin v.s. DPP4I | 0.64[0.51-0.81];0.0002*** | 0.71[0.52-0.98];0.0372* |
| Ertugliflozin v.s. DPP4I | 0.61[0.43-0.87];0.0060** | 0.54[0.32-0.90];0.0192* |

### Subgroup stratification analysis results

As expected, both SGLT2I and DPP4I user above the age of 65 had higher associations with new onset cataracts, though in both the over 65 group and in the under 65 group, DPP4I users had a stronger association that their SGLT2I counterparts in developing cataracts (**Figure 2A**). DPP4I users under the age of 65 years the highest association of new onset diabetic retinopathy than SGLT2I users of all ages and DPP4I users over the age of 65 years, SGLT2I users over the age of 65 had the lowest association (**Figure 2B**).

**Figure 2.**
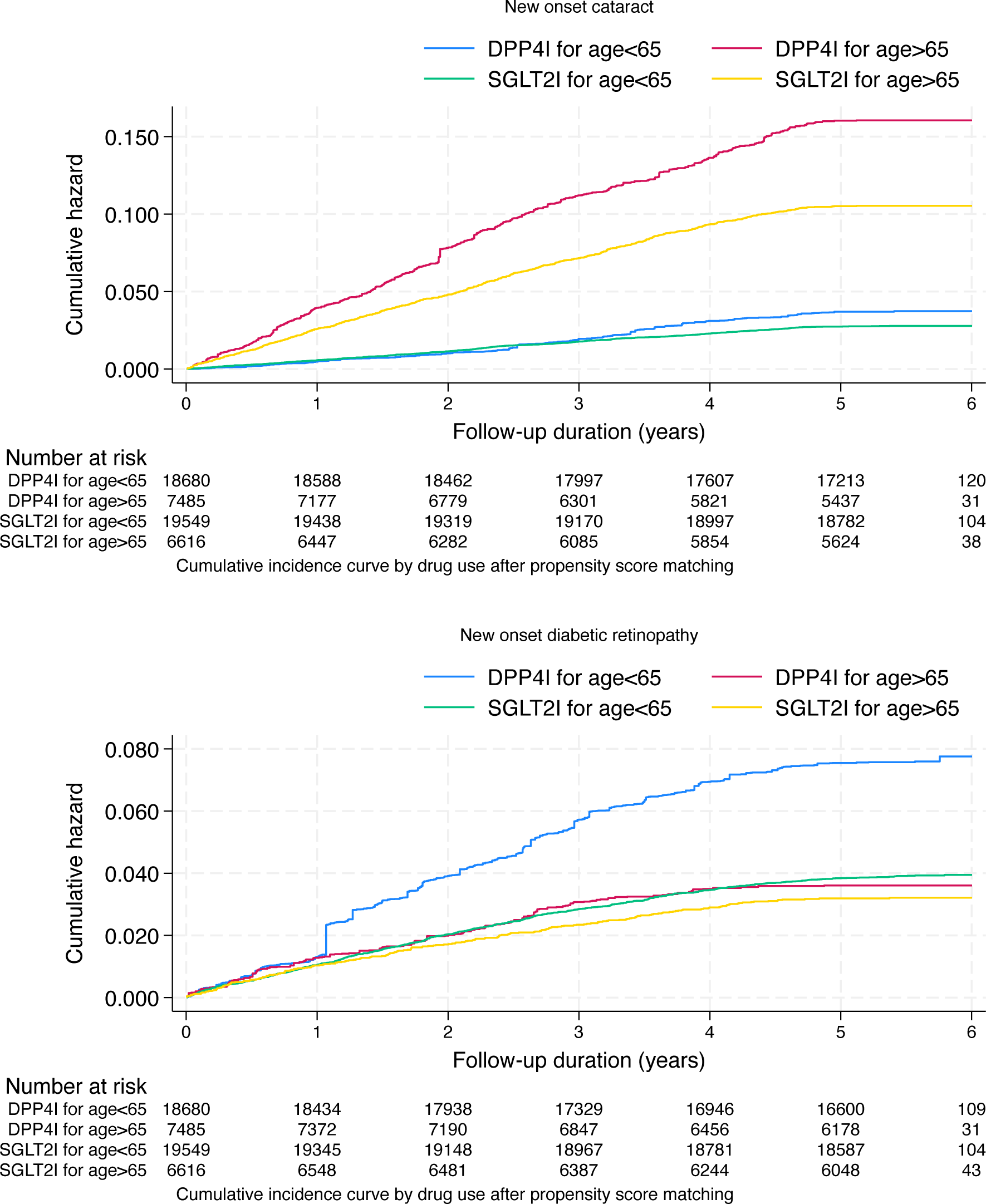
Age-drug specific cumulative incidence curves of new onset cataract and new onset diabetic retinopathy in the matched cohort (1:1) SGLT2I: sodium glucose cotransporter-2 inhibitor; DPP4I: dipeptidyl peptidase-4 inhibitor.

**Figure 3.**
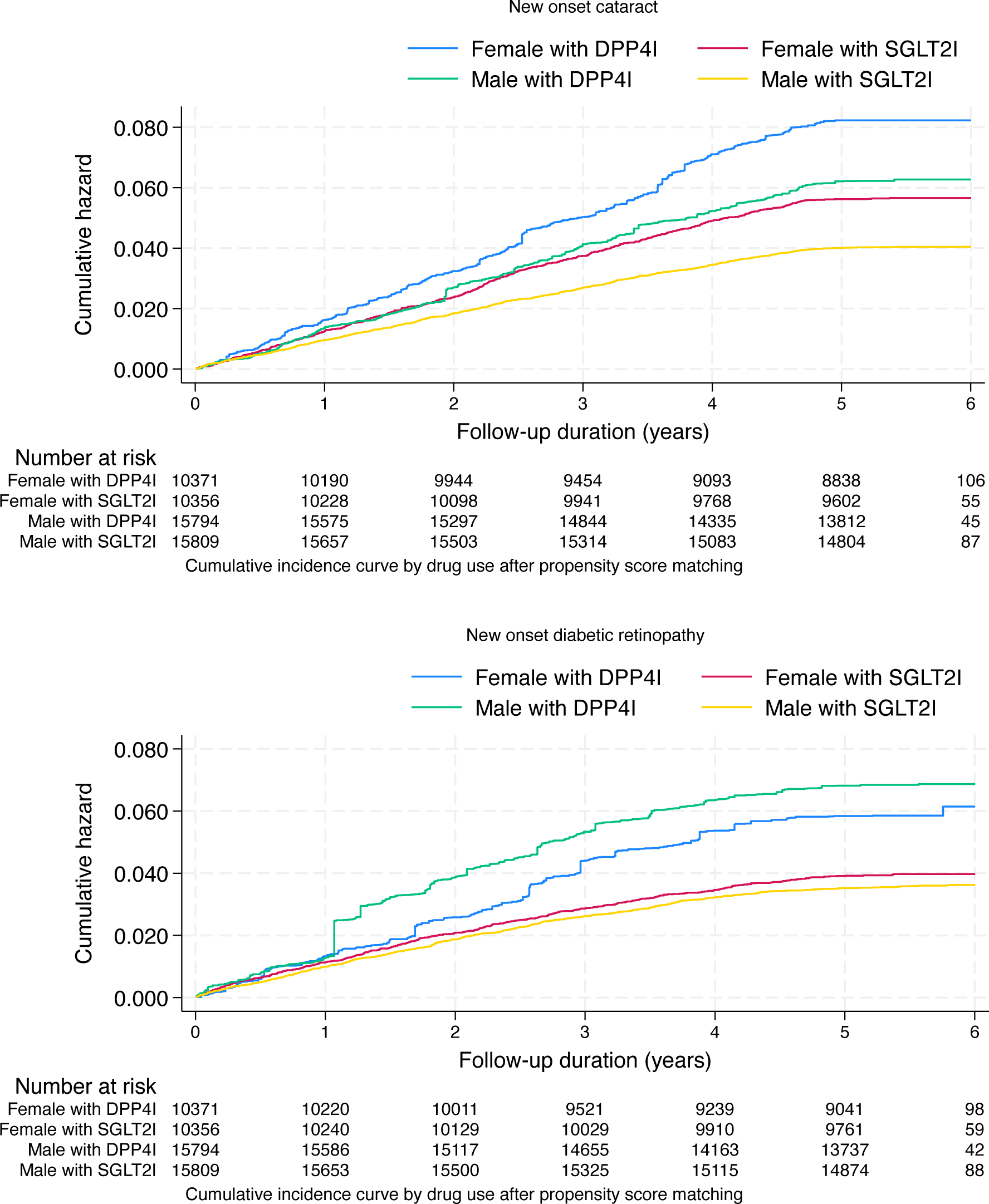
Sex-drug specific cumulative incidence curves of new onset cataract and new onset diabetic retinopathy in the matched cohort (1:1) SGLT2I: sodium glucose cotransporter-2 inhibitor; DPP4I: dipeptidyl peptidase-4 inhibitor.

When stratified by sex, both males and females using DPP4I had a higher association with new onset cataracts compared to SGLT2 users and within the respective DPP4I and SGLT2I groups, females tended to have a higher association with developing cataracts (**Figure 2C**). It was found that males and females using DPP4I had a higher risk of incident diabetic retinopathy than SGLT2I users, but within the DPP4I group, males had a higher association within developing new diabetic retinopathy, whereas in the SGLT2I group women had a slightly higher association with developing new diabetic retinopathy (**Figure 2D**).

## Discussion

This present study found that amongst patients with T2DM, SGLT2I use was statistically significantly associated with lower incidences new onset cataracts and diabetic retinopathy compared to DPP4I use. The number needed to treat was 10.08 (abosolute risk reduction (ARR) 0.099); 10.08 type 2 diabetic patients needed to be treated with SGLT2I versus DPP4I to prevent to the onset of cataracts in one patient (**Supplementary Table 1**). 7.37 (ARR 0.14) type 2 diabetic patients needed to be treated with SGLT2I versus DPP4I to prevent to the onset of diabetic retinopathy in one patient.

Energy derived from glucose maintains transparency in the intraocular lens. Glucose uptake is thought to be either facilitated through various glucose transporter (GLUT) channels or is sodium-dependent through members of the SGLT family (16). In animal studies, long-term hyperglycemia therefore causes an upregulation of GLUT and the development of advanced glycation end products (16). This in turn mediates NADPH oxidation activation, triggering overproduction of reactive oxygen species, which interrupts the normal functionining of lens fibres leading to opacification (16). However, it has been observed that the SGLT2I dapagliflozin inhibits these overproductions, therefore preventing cataract formation (16, 17). Correspondingly, this study found that SGLT2I users had a statistically significantly lower association with new onset cataract (HR: 0.76, 95% CI: [0.67–0.87 P<0.0001) (**Table 4**). This statistically significantly lower association with new onset diabetic retinopathy was present for the common SGLT2I medication dapagliflozin but not canagliflozin, ertugliflozin and empagliflozin (**Table 4**), suggesting that there may be individual SGLT2I molecule differences which provide factors protective agains cataract formation, though further randomized control trials will be needed to fully establish this.

SGLT2 is expressed on the renal epithelial cells lining the proximal convoluted tubule and serves to resorb glucose and sodium from the glomerular filtrate (18). SGLT2I represses this resorption and therefore, promotes increased glycosuria as well as increased sodium and water excretion, thereby reducing blood glucose (18). The benefits of SGLT2i on reducing blood glucose levels and improving cardiovascular outcomes are well-established (19, 20). However, SGLT2I may have a direct effect on diabetic retinopathy as SGLT2 receptors are also found on retinal pericytes (18). When extracellular glucose concentrations rise, SGLT2 receptors are activated, causing an influx of glucose and sodium (18). The overabundance of sodium causes cell swelling, contraction, and eventually pericyte loss, which is the initial step in diabetic retinopathy development (18). This mechanism is supported by a study which found that the SGLTI, ipragliflozin reduces diabetic retinopathy progression in diabetic Torii fatty rats, in a dose-dependent manner (17). This study which found that SGLT2I users had a statistically significantly lower association new onset diabetic retinopathy (HR:0.51, 95% CI: [0.42 – 0.61, P <0.0001) (**Table 4**). In the post-hoc analysis of the Empagliflozin Cardiovascular Outcome Event Trial in Type 2 Diabetes Mellitus Patients (EMPA-REG OUTCOME) trial, there is no significant statistical difference in the incidence of diabetic retinopathy between empagliflozin and placebo users, which is in keeping with this study in that empagliflozin does not seem to be associated with a lower risk of diabetic retinopathy compared to DPP41. Recent studies cross-sectional studies in Taiwan and South Korean had similar findings (21, 22). Specifically, Yen et al found that empagliflozin, dapagliflozin, and canagliflozin were associated with a significantly lower risk of sight-threatening retinopathy than DPP-4i. This study showed that statistically significantly lower association with new onset diabetic retinopathy was present for the common SGLT2I medication dapagliflozin but not canagliflozin, ertugliflozin and empagliflozin (**Table 4**). The differences in findings could be explained by variations in the study protocols, but also largely due to the fact that Yen et al studies sight-threatening retinopathy and this study looks at new-onset diabetic retinopathy, therefore likely taking into account less severe forms of diabetic retinopathy as well. Dziuba et al estimated 20-year microvascular complications in patients with T2DM and found that the addition on dapagliflozin to current treatment was associated with a 9.8% decrease in incident diabetic retinopathy compared with standard care (23). Therefore, recent evidence may suggest that out of all the SGLT2I medications, dapagliflozin may be associated better diabetic retinopathy outcomes, though further work will be needed to establish if a true protective effect is indeed conferred by dapagliflozin. Furthermore, other SGLT2I medications may also be beneficial for other diabetic complications; an anecdotal report found marked regression of diabetic macular oedema after 16 weeks of ipragliflozin (24).

DPP4I is an enzyme which is ubiquitously expressed on the surface of a variety of cells (25). It is an exopeptidase which selectively cleaves N-terminal dipeptides from a variety of substrates, including cytokines, growth factors, neuropeptides, and incretin hormones, rendering them ineffective (25). DPP4I therefore function in diabetes by increasing the presence of incretin hormones including glucagon-like peptide-1 and glucose-dependent insulinotropic polypeptide, which are major regulators of post-prandial insulin secretion, subsequently prolonging the presence of insulin after food intake and supressing glucagon secretion (26). It has been postulated that DPP4I may activate the stromal cell derived factor-1 alpha Src family tyrosine kinase vascular endothelial cadherin signalling pathway which causes retina vascular leakage and therefore worsens diabetic retinopathy (9, 27). Additionally, the Trial Evaluating Cardiovascular Outcomes with Sitagliptin (TECOS) study found diabetic retinopathy events increased by 21.4% in the treatment group versus placebo, though this was not statistically significant (28). However, other studies suggest that DPP4I does not increase the risk of diabetic retinopathy and overall blindness (29, 30). On the other hand, there are other theories that DPP4I may be protective against diabetic retinopathy, supported by animal models (rats) that proposed that DPP4Is including sitagliptin, vildagliptin and may prevent breakdown of the blood retinal barrier, inhibit the overexpression of factors including vascular endothelial growth factor and prevent retinal pericyte loss, all of which contribute to the development and progression of diabetic retinopathy (31–33). Retrospective studies have shown that DPP4I significantly reduced the progression of diabetic retinopathy in T2DM patients when compared to other oral anti-diabetic medications including metformin and sulphonyureas (34, 35).

Currently, the effect of DPP4I on the lens beyond glycaemic control is poorly understood. To the best of our knowledge, this is the first population-based study on the effects of SGLT2I and DPP4I use on the development of cataract amongst patients with T2DM and our results indicate that SGLT2I is associated with lower incidence of cataracts when comparent to DPP4I.

Unsurprisingly, SGLT2I and DPP4I users in older age groups (>65 years) had a higher associations with incident cataracts (**Figure 2A**). Within each age group (over 65 years vs under 65 years) DPP4I users of all ages had a stronger association than their SGLT2I counterparts in developing cataracts (**Figure 2A**). DPP4I users under the age of 65 had a higher risk of new onset diabetic retinopathy. DPP4I users over the ages of 65 and SGLT2I user under the age of 65 had similar sumulative incidence curves with SGLT2I users under the age of 65 ultimately having a slighing higher cumulative risk from 4.5 years onwards (**Figure 2B**). Large cross sectional studies have determined that a younger age of onset of diabetes being an independent risk factor for the development of diabetic retinopathy in a Chinese T2DM population, with the age of 31-45 being the highest risk group, postulating that a that the level of vascular endothelial growth factor in diabetic patients is higher during an earlier age thereby promoting pathological retinal angiogenesis and fibrovascular proliferation during development of diabetic retinopathy (36–39).

When stratifying by sex, both males and females using DPP4I had a higher association with new onset cataracts compared to SGLT2 users and within the respective DPP4I and SGLT2I groups, females tended to have a higher association with developing cataracts (**Figure 2C**). This finding is consistent with evidence that cataracts are more common in women although the exact reason for this is uncertain (40, 41). It was also found that male and females using DPP4I had a higher risk of incident diabetic retinopathy than SGLT2I users, but within each drug group, males had a higher risk than females. Epidemiology studies have recognised that the male gender is an independent risk factor for developing diabetic retinopathy (42). It was also found that males and females using DPP4I had a higher risk of incident diabetic retinopathy than SGLT2I users, but within the DPP4I group, males had a higher association within developing new diabetic retinopathy, whereas in the SGLT2I group women had a slightly higher association with developing new diabetic retinopathy (**Figure 2F**). Epidemiology studies which found that male sex is an independent risk factor of the development of diabetic retinopathy (**Figure 2C**) (42). Potential reasons for this include alterations in ocular blood flow regulation mediated by sex hormones and inflammatory cytokine profiles variations due to sex, in particular oestrogen in women having a vasodilating effect and testosterone in men having a vasoconstrictive effect potentially worsening diabetic retinopathy (42, 43). Therefore it was interesting to note in the SGLT2I group that women had a higher association with new diabetic retinopathy, and possibly coule be explained by a modulating effect of SGLT2I though much further and in depth work will need to be done to investigate this further.

### Limitations

Several limitations were present in this study. First, given its observational nature, there is an inherent information bias due to under-coding, coding errors and missing data. In particular Body Mass Index (BMI) was severely undercoded and therefore unable to be adjusted for in this study. To address this, extensive laboratory results and comorbidities related to cardiovascular disease and HCC were included to infer possible risk variables indirectly. In addition, patient drug compliance could only be assessed indirectly through prescription refills. Secondly, residual and post-baseline confounding may be present despite robust propensity-matching, especially with the unavailability of information on lifestyle factors. We did try to adjust for severe hypoglycaemia and hyperglycaemia using hospitalisations (for any reason) as a proxy measure for this but acknowledge that this is not ideal. Additionally, information on the stage of diabetic retinopathy, progression of diabetic retinopathy, presence or absence of diabetic macular oedema, type of cataract, and complications from both ocular diseases were not available. Furthermore, surgical data for cataract and diabetic retinopathy were not available. Data on visual acuity was also unavailable. Hong Kong does have a diabetic retinopathy screening programme executed by the risk assessment and management programme (RAMP) under the Hong Kong Hospital Authority, but not all patients joined this programme but those who did, would have the relevant ICD codes confirmed if diagnosed with cataract or retinopathy (44). It should be emphasized that our big data approach has limitations and is complementary to other research methods, such as retrospective registry review (45), cross-sectional recruitment from hospitals (46) or the community (47) or clinical trials (48), which have the advantages of obtaining specific data fields that are not routinely collected.

## Conclusions

Among T2DM patients in Hong Kong, SGLT2I use was associated with lower risks of incident cataract and new onset diabetic retinopathy compared to DPP4I use after multiple adjustment models. This study provides data for further evaluation of SGLT2I and DPP4I use in preventing the incidence and progression of cataracts and diabetic retinopathy in a T2DM individuals. This study may also aid clinicians in deciding between SGLT2 and DPP4I if microvascular complications and cataracts are a concern in individual cases.

## Author Declarations

### Ethics approval and consent to participate

This study was approved by the Institutional Review Board of the University of Hong Kong/Hospital Authority Hong Kong West Cluster (HKU/HA HKWC IRB) (UW-20-250) and complied with the Declaration of Helsinki.

### Consent for publication

The authors consent to the publication of manuscript.

### Availability of data and materials

The data that support the findings of this study were provided by the Hong Kong Hospital Authority, but restrictions apply to the availability of these data, which were used under license for the current study, and so are not publicly available. Data are however available from the authors upon reasonable request and with permission of Hong Kong Hospital Authority.

### Authors’ contributions

LYG, JZ: data analysis, data interpretation, statistical analysis, manuscript drafting, critical revision of manuscript OH, TTLL, JMHH, YHAL, CTC, WTW, CC, QPZ, SL: manuscript drafting, critical revision of manuscript GT: study conception, study supervision, project planning, data interpretation, statistical analysis, manuscript drafting, critical revision of manuscript

## Acknowledgements

None.

## Guarantor Statement

All authors approved the final version of the manuscript. GT is the guarantor of this work and, as such, had full access to all the data in the study and takes responsibility for the integrity of the data and the accuracy of the data analysis.

